# Digital Assessment of Real-World Physical Activity in Pulmonary Hypertension: A Systematic Review and Meta-Analysis

**DOI:** 10.64898/2026.05.08.26351469

**Authors:** Simone Brehm, Luana Fiengo Tanaka, Yasir Majeed, Michaela Barnikel, Christine Le Roux, Alessandro Ghiani, Carl-Philipp Jansen, Simon U Jaeger

## Abstract

**Background:** The assessment of daily-life physical activity (DLPA) using wearables in patients with pulmonary hypertension (PH) can provide information on real-world function, potentially enhancing the evaluation of disease progression.

**Research Question:** What is the existing evidence on sensor-based DLPA assessment in patients with PH and its quality?

**Study Design and Methods:** We searched MEDLINE and Embase from inception to January 13, 2026, extracting data on devices, DLPA outcomes, and associations with clinical outcomes. We obtained pooled estimates through random-effects models and assessed evidence quality using a customized tool.

**Results:** We identified 33 studies (29 adult, 4 pediatric) including 1,257 patients mainly with pulmonary arterial hypertension (PAH), followed by chronic thromboembolic PH (CTEPH), and only rarely with PH due to lung diseases and/or hypoxia. Participants were predominantly female, WHO functional class II-III. Most studies investigated step count and time spent in different physical activity levels, but showed substantial heterogeneity in devices and their utilization. The meta-estimate was 4,811 daily steps. A moderate positive correlation was found between daily step count and six-minute walking distance (6MWD) (r=0.59, 95%CI 0.47-0.69); a weak positive correlation was found between time spent in moderate-to-vigorous physical activity and 6MWD (r=0.38; 95% CI 0.26-0.49). Inconsistent wear-time definition, non-wear reporting and temporal misalignment of DLPA may compromise validity and comparability.

**Interpretation:** Wearable-based DLPA assessment in PH is feasible, though high-quality evidence remains scarce. Future research should standardize procedures, terminology, and reporting of DLPA outcomes. Concordance with established measures such as the 6MWD, and their ability to predict clinical outcomes and disease progression need to be demonstrated.

---

Key Words List: Pulmonary hypertension, Accelerometry, Physical activity, Real-world walking activity, Daily-life physical activity, Digital mobility outcomes, Meta-analysis, Systematic review

## Introduction

Pulmonary hypertension (PH) is a rare but severe disease causing a major impact on patients’ daily functioning and quality of life^1^. The ESC/ERS guideline categorizes PH into five distinct groups with differing etiology, hemodynamics, and therapeutic implications^2^: (1) idiopathic pulmonary arterial hypertension, PAH: (2) due to left heart disease; (3) resulting from lung disease; (4) chronic thromboembolic PH (CTEPH), and (5) unclear/multifactorial mechanisms. All forms manifest with exertional dyspnea and may progress to right heart failure^3^.

Given the impact of PH on patients’ functionality, measures of physical capacity and performance are used for risk stratification, prognosis, and treatment decision-making^4^. These physical activity measures include clinical assessments, such as the WHO functional class (FC) and the 6-minute walk test (6MWT)^2^ and patient-reported outcomes based on PH-specific questionnaires, such as CAMPHOR^5^ and EmPHasis-10^6^.

While questionnaires are indispensable for capturing patients’ perspectives, they are often affected by recall bias^7^. Clinical assessments have limited discriminatory validity, especially between WHO-FC II and III^8,9^ and only produce snapshot views of physical capacity.

As a known predictor of morbidity and mortality in PAH^10,11^, the 6-minute walk distance (6MWD) is conventionally incorporated in clinical trials as either the primary endpoint or as part of a composite primary endpoint^12^. However, favorable clinical effects of certain PAH medications are incongruent with changes in the 6MWD^13^. Further limitations of the 6MWT include high within-subject (day-to-day) performance variability and limited availability of a 30m long walkway^14–17^.

Assessment of daily-life physical activity (DLPA) has the potential to look beyond these traditional measures. DLPA outcomes valued most by patients include total walking distance, ability to walk up hills or inclines, sufficient duration of continuous walking episodes, and engaging in moderate to vigorous activity^1^. These can already be captured reliably using wearable devices.

Although DLPA outcomes are increasingly gaining acceptance as endpoints for clinical trials and routine care^18^, their assessment is unstandardized and their validity not yet firmly established for PH. Preliminary evidence suggests that small sample sizes, a lack of clearly pre-defined outcomes, inconsistent recording procedures and clinically meaningful follow-up may limit the ability to draw definite conclusions regarding external validity.

To address this gap, this systematic review and meta-analysis synthesized evidence from the studies assessing DLPA in PH using wearable devices, focusing on reported outcomes and their correlations with established PH-specific clinical outcomes. This evidence will inform the standardization of DLPA assessment and support the integration of DLPA outcomes as validated endpoints in clinical trials and routine PH care.

## Methods

### Data sources and search strategy

This systematic review was conducted according to PRISMA guidelines^19^ and registered with PROSPERO (CRD42024565518). The search on PubMed and Embase was performed from inception to February 7, 2025, and updated on January 13, 2026. The search strategy comprised terms for PH, physical activity, and wearable devices (Supplementary file: Table 3). Additionally, the reference lists of retrieved studies were manually screened to identify relevant studies.

### Study selection and data extraction

The retrieved articles were exported to EndNote^TM^, duplicates were removed, and two independent investigators (S.B. and Y.M.) assessed them. Articles were initially selected based on title and abstract, followed by full text for final inclusion (Figure 1). A third investigator (S.U.J.) resolved any discrepancies. Data extraction was performed using a standardized collection sheet. When multiple publications reported data from the same population, a primary publication was identified to represent the study, while data from all related publications were extracted and combined to comprehensively capture DMOs, methodological characteristics, and clinical parameters.

**Figure 1.**
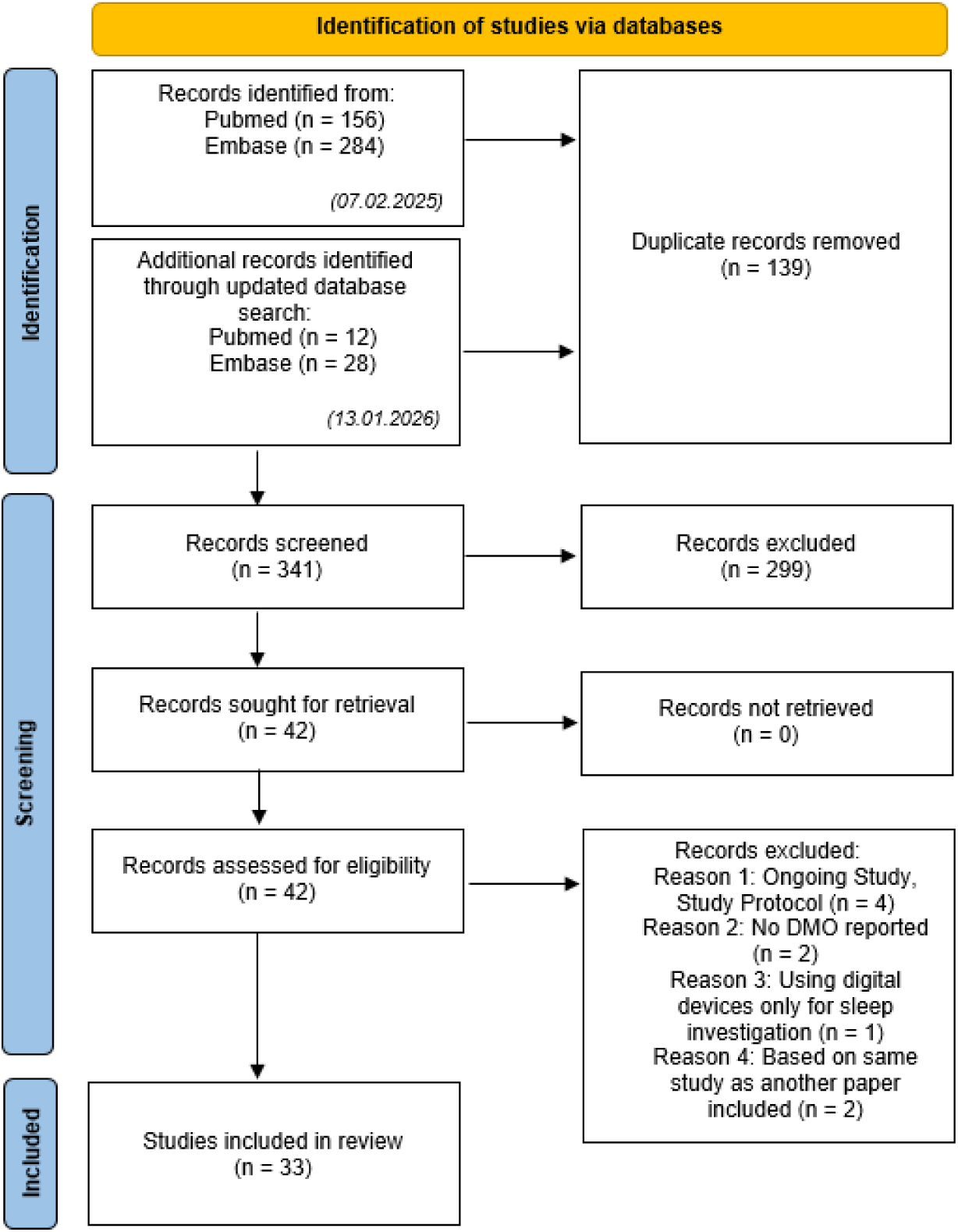
Flowchart.

### Inclusion/Exclusion criteria

Studies meeting the following eligibility criteria were included: peer-reviewed original research (intervention, cohort, and cross-sectional studies) in German or English, reporting at least one DLPA outcome measured with a wearable device in humans with PH. Exclusion criteria were: commentaries, conference abstracts, review articles, study protocols, studies assessing physical activity exclusively in experimental or clinical settings, as well as those focusing solely on sleep behavior without examining everyday activity, publications reporting solely heart rate without providing any measure of physical activity, and those based only on self-reported physical activity. No age restrictions were applied.

### Evaluation of the methodological conduct and reporting of studies

We applied a customized tool to evaluate the quality of wearable-based DMOs reported by studies. Seven criteria were defined^20,21^:

1. Sufficient information on device type and model is provided.
2. Multiple digital parameters describing DLPA are reported.
3. Measures of central tendency and variability of DLPA outcomes are provided.
4. Minimal wear time is reported to define “valid days”.
5. Mean/median wear time is reported.
6. Actual device wear time is at least 3 valid days with ≥10 h wear time.
7. Device was worn continuously throughout the study period.

Each criterion was rated as adequate, inadequate, or not reported. “Not reported” was used when essential methodological details were missing. “Inadequate” was applied when information was incomplete or insufficiently explained (Table 1).

**Table 1.**
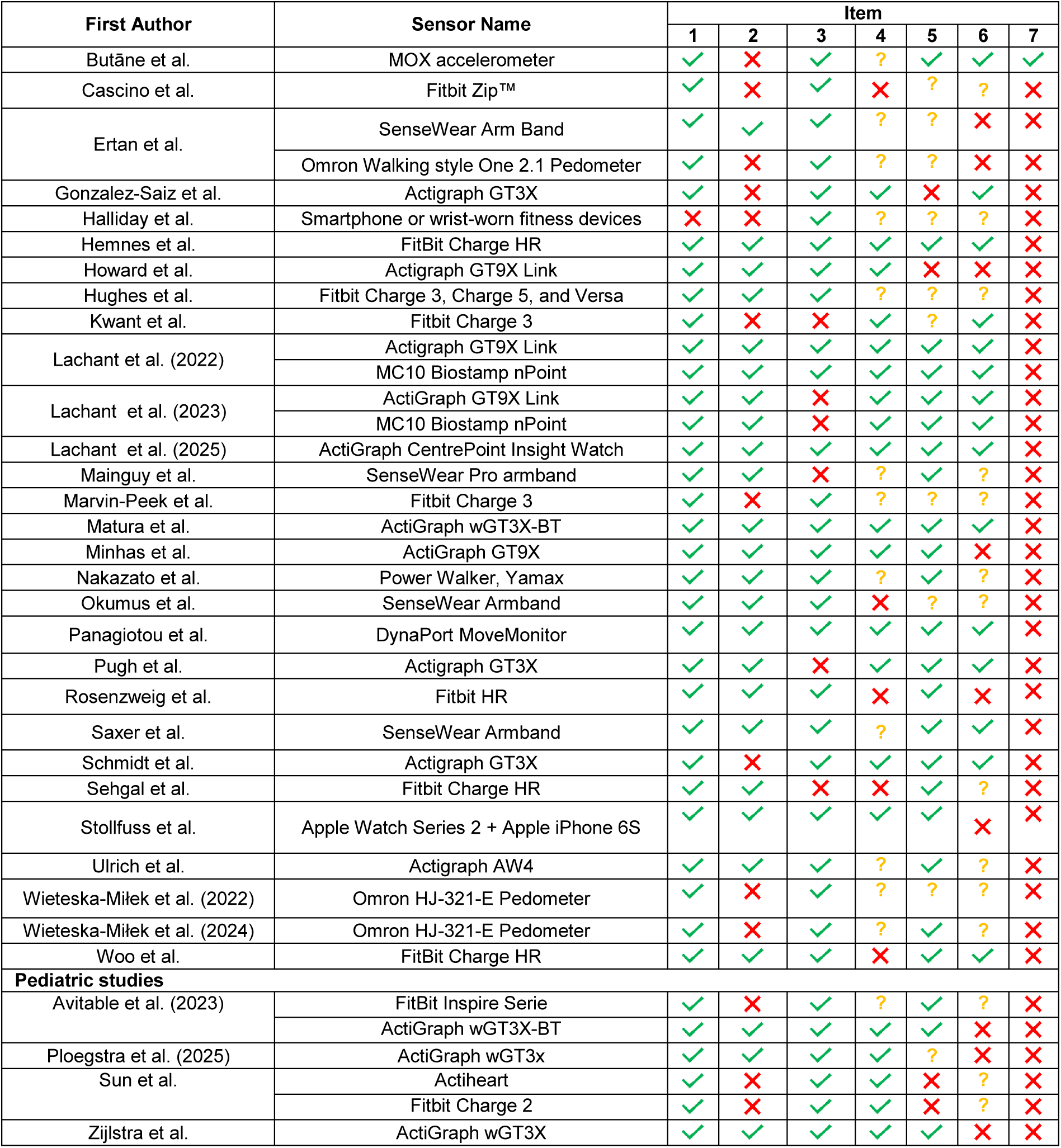
Quality assessment. 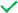 Adequate, 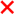 Inadequate, 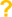 Not reported; Item 1: Sufficient information on the device type and model is provided, Item 2: The study reports multiple digital parameters describing daily-life physical activity, Item 3: Reported DMOs include measures of central tendency and variability, Item 4: A minimal wear duration is specified to define “valid days”, Item 5: A mean/median wear time is reported, Item 6: The actual device wear time is at least 3 valid days with ≥ 10 h wearing time, Item 7: The device is worn continuously (i.e. non-removable) throughout the study period.

| First Author | Sensor Name | Item |  |  |  |  |  |  |
| --- | --- | --- | --- | --- | --- | --- | --- | --- |
|  |  | 1 | 2 | 3 | 4 | 5 | 6 | 7 |
| Butāne et al. | MOX accelerometer | ✓ | ✗ | ✓ | ? | ✓ | ✓ | ✓ |
| Cascino et al. | Fitbit Zip™ | ✓ | ✗ | ✓ | ✗ | ? | ? | ✗ |
| Ertan et al. | SenseWear Arm Band | ✓ | ✓ | ✓ | ? | ? | ✗ | ✗ |
|  | Omron Walking style One 2.1 Pedometer | ✓ | ✗ | ✓ | ? | ? | ✗ | ✗ |
| Gonzalez-Saiz et al. | Actigraph GT3X | ✓ | ✗ | ✓ | ✓ | ✗ | ✓ | ✗ |
| Halliday et al. | Smartphone or wrist-worn fitness devices | ✗ | ✗ | ✓ | ? | ? | ? | ✗ |
| Hemnes et al. | FitBit Charge HR | ✓ | ✓ | ✓ | ✓ | ✓ | ✓ | ✗ |
| Howard et al. | Actigraph GT9X Link | ✓ | ✓ | ✓ | ✓ | ✗ | ✗ | ✗ |
| Hughes et al. | Fitbit Charge 3, Charge 5, and Versa | ✓ | ✓ | ✓ | ? | ? | ? | ✗ |
| Kwant et al. | Fitbit Charge 3 | ✓ | ✗ | ✗ | ✓ | ? | ✓ | ✗ |
| Lachant et al. (2022) | Actigraph GT9X Link | ✓ | ✓ | ✓ | ✓ | ✓ | ✓ | ✗ |
|  | MC10 Biostamp nPoint | ✓ | ✓ | ✓ | ✓ | ✓ | ✓ | ✗ |
| Lachant et al. (2023) | ActiGraph GT9X Link | ✓ | ✓ | ✗ | ✓ | ✓ | ✓ | ✗ |
|  | MC10 Biostamp nPoint | ✓ | ✓ | ✗ | ✓ | ✓ | ✓ | ✗ |
| Lachant et al. (2025) | ActiGraph CentrePoint Insight Watch | ✓ | ✓ | ✓ | ✓ | ✓ | ✓ | ✗ |
| Mainguy et al. | SenseWear Pro armband | ✓ | ✓ | ✗ | ? | ✓ | ? | ✗ |
| Marvin-Peek et al. | Fitbit Charge 3 | ✓ | ✗ | ✓ | ? | ? | ? | ✗ |
| Matura et al. | ActiGraph wGT3X-BT | ✓ | ✓ | ✓ | ✓ | ✓ | ✓ | ✗ |
| Minhas et al. | ActiGraph GT9X | ✓ | ✓ | ✓ | ✓ | ✓ | ✗ | ✗ |
| Nakazato et al. | Power Walker, Yamax | ✓ | ✓ | ✓ | ? | ✓ | ? | ✗ |
| Okumus et al. | SenseWear Armband | ✓ | ✓ | ✓ | ✗ | ? | ? | ✗ |
| Panagiotou et al. | DynaPort MoveMonitor | ✓ | ✓ | ✓ | ✓ | ✓ | ✓ | ✗ |
| Pugh et al. | Actigraph GT3X | ✓ | ✓ | ✗ | ✓ | ✓ | ✓ | ✗ |
| Rosenzweig et al. | Fitbit HR | ✓ | ✓ | ✓ | ✗ | ✓ | ✗ | ✗ |
| Saxer et al. | SenseWear Armband | ✓ | ✓ | ✓ | ? | ✓ | ✓ | ✗ |
| Schmidt et al. | Actigraph GT3X | ✓ | ✗ | ✓ | ✓ | ✓ | ✓ | ✗ |
| Sehgal et al. | Fitbit Charge HR | ✓ | ✓ | ✗ | ✗ | ✓ | ? | ✗ |
| Stollfuss et al. | Apple Watch Series 2 + Apple iPhone 6S | ✓ | ✓ | ✓ | ✓ | ✓ | ✗ | ✗ |
| Ulrich et al. | Actigraph AW4 | ✓ | ✓ | ✓ | ? | ✓ | ? | ✗ |
| Wieteska-Milek et al. (2022) | Omron HJ-321-E Pedometer | ✓ | ✗ | ✓ | ? | ? | ? | ✗ |
| Wieteska-Milek et al. (2024) | Omron HJ-321-E Pedometer | ✓ | ✗ | ✓ | ? | ✓ | ? | ✗ |
| Woo et al. | FitBit Charge HR | ✓ | ✓ | ✓ | ✗ | ✓ | ✓ | ✗ |
| **Pediatric studies** |  |  |  |  |  |  |  |  |
| Avitable et al. (2023) | FitBit Inspire Serie | ✓ | ✗ | ✓ | ? | ✓ | ? | ✗ |
|  | ActiGraph wGT3X-BT | ✓ | ✓ | ✓ | ✓ | ✓ | ✗ | ✗ |
| Ploegstra et al. (2025) | ActiGraph wGT3x | ✓ | ✓ | ✓ | ✓ | ? | ✗ | ✗ |
| Sun et al. | Actiheart | ✓ | ✗ | ✓ | ✓ | ✗ | ? | ✗ |
|  | Fitbit Charge 2 | ✓ | ✗ | ✓ | ✓ | ✗ | ? | ✗ |
| Zijlstra et al. | ActiGraph wGT3X | ✓ | ✓ | ✓ | ✓ | ✓ | ✗ | ✗ |

### Calculation of pooled estimates for DMOs and correlations

We only performed data pooling (DLPA outcomes and correlations) when estimates were available from at least three studies. To allow for comparability between cross-sectional and longitudinal studies, only baseline estimates were used. For studies reporting medians and interquartile ranges, means were estimated using the method of Luo et al.^22^ and standard deviations (SD) were estimated using the method of Wan et al., assuming approximate normality^23^.

We pooled estimates using a random-effects model due to important differences in study populations and study designs. We excluded pediatric studies from pooling due to their low comparability with the adult population. All analyses were conducted using R version 4.4.2 with the metafor package (version 4.8-0).

## Results

### Study characteristics and patient population

Thirty-three studies published between November 2011 and January 2026 were included. Among these, 26 were observational (19 cross-sectional, 7 cohort), and seven were interventional studies. Most studies were conducted in the USA (17)^24–37^, with a multinational investigation including sites in the USA and Canada^38^ and a large multi-country trial^39^. Further work originated from Europe, Canada, South America, and Turkey.

The majority of the studies (n=24) covered exclusively patients with PAH. Seven studies included both PAH and CTEPH patients; one pediatric study enrolled PAH and PH Group 3 patients, while one study focused solely on CTEPH patients. Most studies enrolled adults (27), with two studies including adolescents aged 13 years and older^33,35^ and four were dedicated to pediatric populations^24,38,40,41^.

Across all studies, 1,257 subjects (87% adults) with PH were evaluated. The mean ages were 10 years (±10.4 years) in the pediatric and 51.8 years (±17.4 years) in studies with adolescents/adults. Patients were predominantly female (75.1%), and most had moderate functional impairment (WHO-FC II-III).

Only 17 studies provided catheterization data, and fewer (9) linked these hemodynamic measurements to DLPA outcomes. No study described follow-up hemodynamic data. 6MWD and WHO-FC were the most commonly reported clinical outcomes. Additional parameters included echocardiographic measurements, N-terminal pro-B-type natriuretic peptide (NT-proBNP/BNP) levels, spirometry, and cardiopulmonary exercise testing (CPET) results collected primarily at baseline (Table 2).

**Table 2.** Study and population characteristics; only the number of patients that were actually analyzed by the authors were included; *Displayed numbers indicate how many studies reported the DLPA outcome or clinical parameter; Studies mentioning RHC solely as a diagnostic requirement were not counted as reporting RHC data;: 6MWD: six-minute walk distance, BNP: brain natriuretic peptide, CPET: Cardiopulmonary exercise testing, CTEPH: Chronic Thromboembolic Pulmonary Hypertension, FU: Follow-up, IQR: interquartile range, NT-proBNP: N-terminal prohormone of brain natriuretic peptide, PAH: pulmonary arterial hypertension, PH: pulmonary hypertension, WHO-FC: WHO Functional Class

| Studies (n = 33) |  |  |  |
| --- | --- | --- | --- |
| Study type | Observational | Cross-sectional, n | 19 |
|  |  | Cohort, n | 7 |
|  | Interventional, n |  | 7 |
| Age category | Pediatric, n |  | 4 |
|  | Adults ≥ 18 years, n |  | 27 |
|  | Adolescent and Adults ≥ 13 years, n |  | 2 |
| Sample size, median (IQR) |  |  | 25 (20-43) |
| PH Population (n = 1,257) |  |  |  |
|  |  | Adolescent and Adults<br>(n = 1,094) | Children<br>(n=163) |
| PH classification, n (%) | PAH | 993 (90.8) | 155 (95.1) |
|  | Group3 | - | 8 (4.9) |
|  | CTEPH | 101 (9.2) | - |
| Gender, n (%) | Female | 822 (75.1) | 97 (59.5) |
|  | Male | 252 (23.0) | 66 (40.5) |
|  | Missing participants, n | 20 (1.8) | - |
| Age, mean of study means<br>(SD of mean) |  | 51.8 (17.4) | 10.0 (4.3) |
|  | Missing participants, n | 38 | - |
| NYHA/WHO-FC, n (%) | I | 110 (10.1) | 21 (12.9) |
|  | II | 572 (52.3) | 100 (61.3) |
|  | III | 270 (24.7) | 40 (24.5) |
|  | IV | 13 (1.2) | 2 (1.2) |
|  | Missing | 129 (11.8) | - |
| Amount of studies that reported DLPA outcomes* |  |  |  |
|  | Baseline |  | FU |
| Steps per day | 25 |  | 10 |
| Cadence (steps/min) | 1 |  | 1 |
| Total activity time (active/inactive) | 9 |  | 2 |
| Time spent at different PA/intensity levels (e.g. (non)sedentary, light, standing, moderate, mid to vigorous, (very) vigorous) | 18 |  | 8 |
| (Total / active) energy expenditure | 3 |  | 1 |
| Metabolic equivalent of task | 3 |  | 1 |
| Vector magnitude counts/min | 9 |  | 3 |
| Distance walked per day | 1 |  | 1 |
| Number of standing-up events per day | 1 |  | 1 |
| Walking intensity (m/s<sup>2</sup>) | 1 |  | - |
| Walking bouts (number, duration, distance) | 1 |  | 1 |
| Amount of studies that reported clinical outcomes* |  |  |  |
|  | Baseline |  | FU |
| Right heart catheterization | 17 |  | - |
| Echocardiography | 14 |  | 5 |
| NYHA/ WHO-FC | 32 |  | 4 |
| 6MWD | 31 |  | 11 |
| CPET | 4 |  | 2 |
| Spirometry | 3 |  | - |
| NT-proBNP/ BNP | 18 |  | 8 |

### Devices, wear location, duration, and time

Several digital devices were used to measure DLPA: various ActiGraph^TM^ models, Fitbit devices, pedometers, SenseWear ArmBands, the MC10 Biostamp nPoint, Actiwatch model AW4, Actiheart, and others. The devices were worn either at wrist, arm, chest, back, waist, hip, thigh, or leg (Supplementary file: Figure 4). Five studies used multiple devices simultaneously per patient, collecting complementary physiological data^42,43^ or directly comparing device performance^24,28,37^. One study found no difference between devices when comparing step counts^24^ whereas others did^28,37^. Three studies pooled DMOs although the data was captured by different devices^26,31,36^.

Total device wear duration varied across studies (Figure 2). Most studies (n=16)^28,30–32,35,37,39,40,44–50^ planned a measurement period of seven consecutive days, while six studies^24,25,27,29,43,51^ targeted 14 days. Other studies reported measurement periods of several weeks or even months^26,27,33,34,36,39,52–55^. Fourteen studies included follow-up data, with varying measurement designs: five conducted continuous measurements throughout the study period, using either the first and last available measurement points as baseline and follow-up or reporting only changes from baseline without explicitly defining a distinct follow-up time point^27,33,34,39,53^, respectively, whereas the others implemented two or three separate measurement periods, explicitly defining baseline and follow-up periods^28,30,36,41,42,44,52,55,56^.

**Figure 2.**
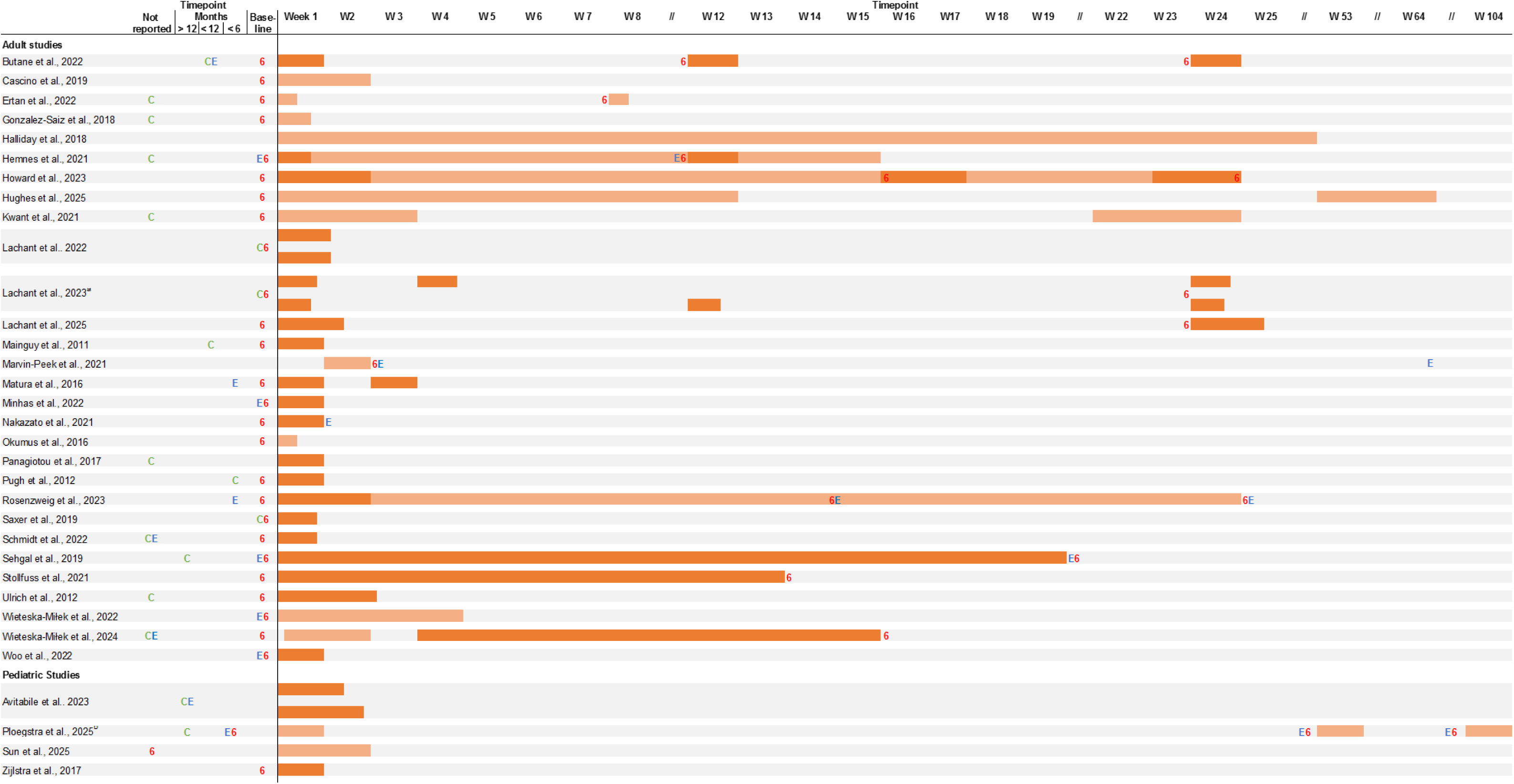
Study timelines of weartime of the activity measurements; Dark orange timeline bars represent the actual reported wear time (mean or median); Timeline bars are colored light orange when actual wear time was not reported, so they represent the intended wear time per protocol; C = Right Heart Catheterization, E = Echocardiography, 6 = 6-Minute Walk Test; ^a^There were only two monitoring periods per participant. All patients wore both devices, but patients were analysed in two groups. The second monitoring period was performed within 4 weeks for the “stable group” and at 3 months for the “treatment intensification group”^28^ ^b^Participants completed one to three accelerometry assessments; 6-minute walk test and echocardiographic data were derived from routine care, were not available at all time points for every participant, and were matched to accelerometry within ±3 months^41^

**Figure 3.**
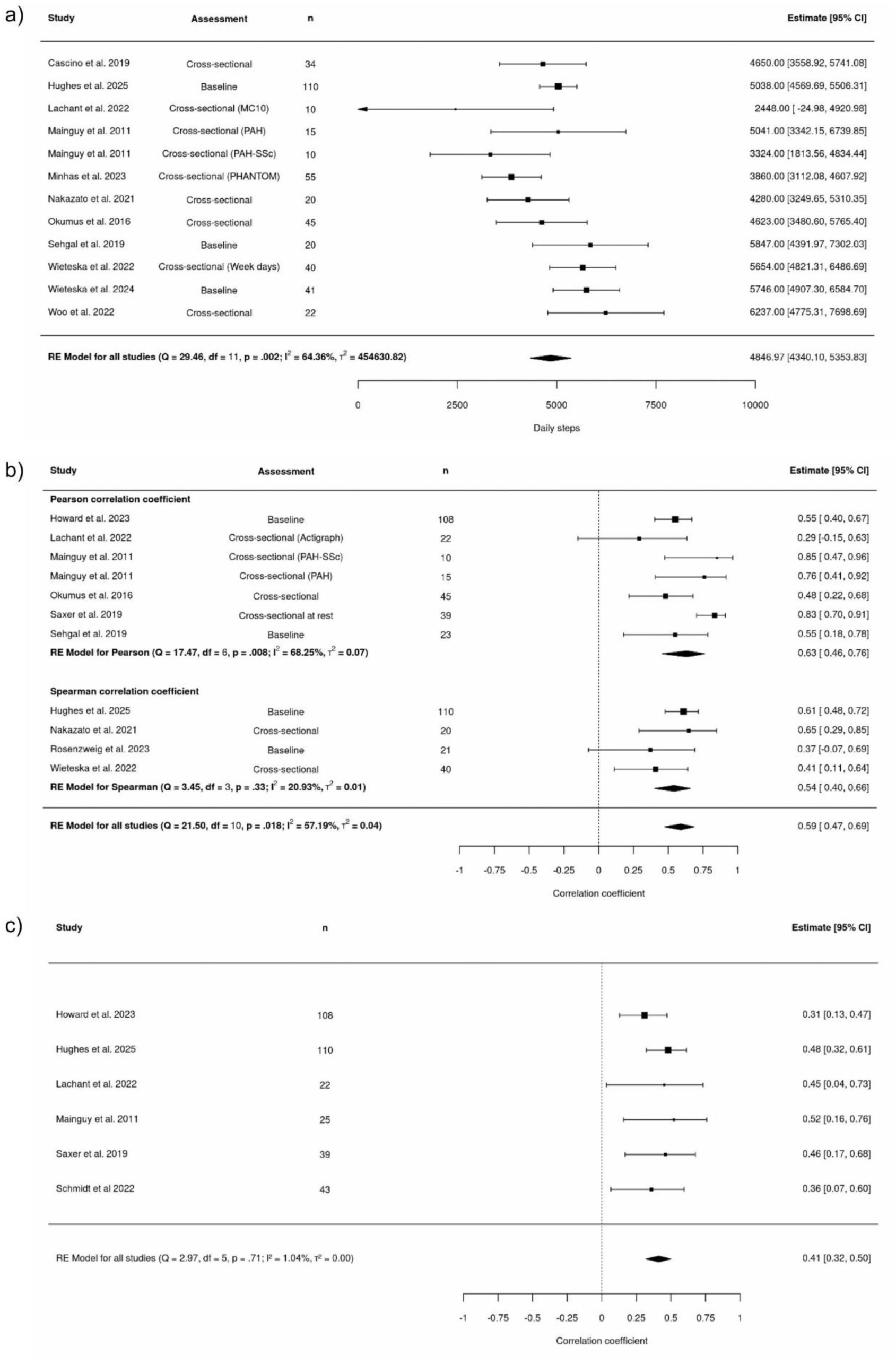
Pooled estimate of a) daily step count based on eleven studies; b) correlation between daily step count and 6-minute walking distance based on eleven studies; c) correlation between time spent at moderate-to-vigorous physical activity and 6-minute walking distance based on six studies; PAH: Pulmonary arterial hypertension, SSc: Systemic sclerosis, RE: Random-effects, CI: Confidence interval, df: degrees of freedom.

For reliable measurement and comparability, actual wear time varied considerably. A valid wear time (total days and hours per day) was defined only in 15 studies, ranging from >2h/day to >20h/day, with a minimum duration of three to seven days. Apart from one study^44^, all studies used a protocol in which the device was not worn continuously throughout the assessment period, thereby increasing the likelihood of non-wear episodes and incomplete data.

### DLPA Outcomes

The digital assessment of DLPA showed substantial methodological heterogeneity, with diverse outcomes being investigated (Table 1). Daily steps were the most consistently reported DLPA outcome cross-sectionally (n=25) and longitudinally (n=10). Lachant et al. additionally analyzed peak steps^28,56^. Beyond steps, studies captured a broad spectrum of activity-related metrics, including raw accelerometer outputs such as vector magnitude counts, overall activity time, and time spent in different physical activity intensity categories. Intensity is often categorized into sedentary versus non-sedentary PA. In addition, light or low, moderate, moderate-to-vigorous, and vigorous or high physical activity are reported as minutes per day or percent of the total awake time. Intensity is also frequently reported (n=7) as device-specific arbitrary value based on vector magnitude counts. Metabolic equivalents (METs), or activity intensity categories based on MET values, were also commonly used to quantify PA intensity. Energy expenditure, standing-up events, daily walking distance, or walking intensity were rarely reported. In the TRACE trial^39,57^, walking activity was further characterized in more depth, including the number of walking bouts (episodes), bout duration (minutes), and bout distance (steps) with additional analyses of steps (steps/minute).

### Associations between DLPA and clinical outcomes

#### 6MWD

The correlation between DLPA outcomes and 6MWD was reported in 15 studies^30–34,37,39,43,46–50,58,59^. In all of those studies, at least one DLPA outcome was positively associated with higher 6MWD. Ten of the 11 studies reporting daily step counts^31,33,34,37,39,43,46,47,49,58,59^ found a statistically significant positive association^37^.

#### WHO-FC

All five studies assessing the association between DLPA outcomes and WHO-FC reported higher FC with lower activity levels^29,32,34,37,50^.

#### Echocardiography

The tricuspid annular plane systolic excursion, a measure of right ventricular systolic function, was most commonly reported^29,31,34,35^, but only Minhas et al. showed a significant correlation with DLPA, with higher values linked to increased vector magnitude counts and a trend toward higher daily step counts.

#### Hemodynamic parameters from RHC

Nine studies collected hemodynamic parameters from RHC for association with DLPA ouctomes^28,32,34,37,45,46,49–51^ where overall, no consistent or robust associations were observed. No significant associations were found for right atrial pressure^32,34,46^ and pulmonary capillary wedge pressure^34,49^. For mean pulmonary arterial pressure, most studies also reported no significant associations with DLPA^32,34,49,50^ except one^45^ that observed a weak but significant inverse correlation with moderate PA. Pulmonary vascular resistance was found to correlate negatively with energy expenditure at maximum exercise, but not at rest^49^, yet others reported no significant association between pulmonary vascular resistance and various DLPA outcomes^34,50^. In contrast, time spent in bed was significantly associated with pulmonary vascular resistance^51^, and daily activity decreased with increasing hemodynamic disease severity. Further, a moderate correlation between cardiac output and energy expenditure during exercise, but not with step count, was found^49^, indicating outcome-dependent associations within the same hemodynamic framework.

#### Patient-reported outcomes

Various PH- or heart failure-specific instruments were employed to assess PH burden. These included emPHasis 10^31,34^, CAMPHOR^30,50^, Manchester respiratory activities of daily living questionnaire^47^ and Minnesota living with heart failure questionnaire^58^. While lower emPHasis 10 scores (representing less impairment) were reported with increasing activity^31^, there was no association between emPHasis 10 and any DLPA outcomes^37^. Lower scores on the CAMPHOR symptoms scale correlated with higher activity in the moderate-to-vigorous range^50^ and with percentage of activity bouts^30^. Matura et al. showed an association of less energy in the energy subscale of the symptoms scale with a lower percentage of episodes of continuous activity above predefined vector magnitude levels^30^.

#### Cardiopulmonary exercise testing (CPET)

Four studies examined the association between CPET and DLPA coutcomes with inconsistent findings. One study observed that more active patients demonstrated better ventilatory efficiency^48^, whereas another reported that patients with higher activity levels were more likely to meet established thresholds in CPET parameters with prognostic significance^45^. Woo et al. reported that accelerometry-based activity showed no meaningful correlation with performance on CPET performance^35^. Rosenzweig et al. did not find a significant association at baseline and follow-up between daily steps and key parameters of CPET^33^.

#### Spirometry

Only three studies provided spirometry data^44,47,48^, and none showed clear associations with DLPA outcomes, except one that observed a significant positive association between transfer factor for carbon monoxide %predicted and daily walking intensity, while other spirometry indices were unrelated to activity^48^.

#### Laboratory markers of heart failure

Four studies examined the association between DLPA outcomes and NT-pro-BNP levels^34,37,48,50^; two studies found a statistically significant correlation between lower NT-pro-BNP and higher activity time or walking intensity, respectively^37,48^.

#### Risk prediction tools

Lachant et al. reported a negative correlation between total activity time and the REVEAL 2.0 risk score^37^. Marvin-Peek et al. reported lower hospitalization rates with increasing daily steps but no association between DLPA outcomes and escalation of the therapeutic regimen^29^.

### Pooled estimates

Eleven studies reported daily steps for 412 patients. The pooled estimate was 4,847 daily steps (95%CI 4,340-5,354 daily steps) with considerable heterogeneity across studies (𝐼^2^= 64.4%). Ten studies reported the correlation between daily steps and 6MWD, covering 343 patients. We detected a moderate positive correlation between daily steps and 6MWD (r = 0.59, 95%CI 0.47-0.69) with significant heterogeneity across studies (𝐼^2^ = 57.2%). The association between time spent in MVPA and 6MWD was 0.41 (95%CI 0.32-0.50) with low study heterogeneity (𝐼^2^ = 1.0%).

## Discussion

This manuscript aimed to synthesize evidence from 33 studies that assessed DLPA using wearable sensors in patients with PH, focusing on correlations between sensor-based DLPA outcomes and established clinical outcomes.

The majority of studies reported cross-sectional data from small adult populations with PAH, using convenience sampling from a single center. Only 11 studies incorporated a follow-up measurement of DLPA, hereby enabling assessment of responsiveness of respective outcomes (sensitivity to change). Our pooled estimates highlighted low numbers of daily steps, a low percentage of time spent at moderate activity levels, and increased sedentary time. At the same time, inconsistent definitions of wear duration, reporting of non-wear and lack of temporal alignments of DLPA with clinical assessments introduces bias, contributes to between-study heterogeneity, and currently limits the acceptance of sensor-based DLPA outcomes in clinical trials and routine practice.

### Daily steps

Daily steps was the most commonly reported DLPA outcome. The pooled estimate in adult PAH/CTEPH patients from this study, of which >70% were in WHO-FC II and III, was 4,847 steps/day. This is comparable to COPD patients with increased symptoms but low exacerbation risk (GOLD B), who walk around 4,850 steps/day^60^. More generally, a recent meta-analysis of 15 international cohorts with 47,471 participants showed that, in a sample of individuals <60 years of age comparable to this analysis, 4,900 daily steps fall into the lowest quartile. Such a low number of daily steps is significantly associated with an increase in all-cause mortality^61^. Prognostic value of daily steps has also specifically been reported for PAH showing that fewer daily steps are associated with worse transplant-free survival and shorter time to PAH-related hospitalization^62^. Marvin-Peek et al. report that walking more than 5,500 steps per day significantly decreased the risk of PAH-related hospitalization within a median follow-up time of 2.2 years^29^.

The association between daily steps and clinical outcomes, laboratory, hemodynamic, and imaging parameters, as well as patient-reported outcomes, was mostly statistically insignificant. The most consistent correlation was between daily steps and 6MWD, with pooled estimates of Pearson’s *r* of 0.63 or Spearman *ρ* = 0.54. This shows that the 6MWT is a useful component of PH risk stratification and is therefore relevant to clinical decision making^63^. For daily steps and WHO-FC, another important clinical outcome, only two studies reported a significant negative correlation: fewer daily steps were associated with higher WHO-FC^29,46^. Considering patient-reported outcomes, no clear picture emerged. Although NT-proBNP levels are part of routine assessments and have prognostic value^2^, only one study reported a significant correlation. Similarly, across all studies, echocardiography findings or hemodynamic parameters showed no clear relationship with daily steps.

Dails steps capture clinically meaningful physical limitations in PAH/CTEPH, as activity levels fall into ranges associated with increased mortality across other diseases. However, limited and inconsistent correlations with a broader range of important clinical outcomes highlight that a broader range of outcomes is necessary to sufficiently reflect DLPA in PAH.

### Physical activity intensity levels

In acknowledgement of limitations inherent to a single bulk measure like daily steps, in most studies, DLPA is described using additional parameters since proprietary analysis algorithms of the accelerometers and gyroscopes readily provide a variety of these.

The ability to perform tasks with a moderate activity intensity such as brisk walking, gardening, or carrying groceries^64^ is particularly important for PH patients’ quality of life^1^. We note that physical activity intensity levels are stratified quite differently across studies, and in the absence of clear frameworks, remain dependent on the device, investigator’s choice and may be data-driven.

As moderate-to vigorous PA is equivalent to 6MWD of 400-500 m, a correlation can be expected, and indeed five studies found a significant positive correlation of moderate-to-vigorous PA (>3 METs) with the 6MWD (meta-estimate at studies’ baseline was 0.38). Such moderate correlations have also been described in other lung diseases, e.g., in COPD^65,66^ or lung transplant candidates with interstitial lung disease^67^.

Significant correlations between intensity levels and WHO-FC have been reported in five studies. For shorter time spent in moderate-to vigorous PA, two studies reported a higher WHO-FC^37,46,50^. The heterogeneity of the intensity parameters reported precluded calculation of a meta-estimate.

Similar to daily steps, both significant and non-significant correlations between PA intensity levels with patient-reported outcomes have been reported. However, due to the heterogeneity of DLPA outcomes and questionnaires used across studies, it was not possible to synthesize overall specific summary measures.

### Temporal alignment of the assessments

An externally valid correlation requires temporal alignment of the assessments. The 6MWD is a regular part of routine visits in most PAH centers, and patient-reported outcomes can be administered easily around the time of DPLA assessment. The orchestration of RHC or echocardiography with DLPA assessment comes with greater logistic challenges. Additionally, as an invasive procedure, RHC cannot be performed solely for the purpose of validating DLPA outcomes and echocardiography remains resource-intensive as well. As a result, evidence regarding correlations of DLPA parameters with hemodynamic or imaging parameters was sparse and not robust. In the absence of documentation of RHC or echocardiography timing and given the frequent temporal misalignment (e.g. >6 months) across studies, the observed associations should be regarded as hypothesis-generating only.

### Technical aspects and validity of measurements

Both research-grade and commercially available devices have been used to measure DLPA. The placement of these devices varies, with options including the hip, wrist, upper arm, thigh, chest and back. Our findings align with those reported in a narrative review of 16 PAH / physical activity studies^68^, which was published several months after our systematic review was registered on PROSPERO. There is a notable lack of standardization across instrumentation, data interpretation methodologies, sensor placement, and software algorithms. Variations in outcomes may therefore be attributed to these differences. These conclusions mirror the ones drawn from our systematic analysis: standardized measurement protocols and further research are required to ascertain the most suitable device and measurement method for PAH cohorts.

The validity and interpretation of DLPA outcomes rests not only on an accurate technical measurement and sensor wear location, but on the actual wear time as well^69^. While no uniform definition of minimum wear time exists, ≥10h/day on ≥4 days^70^ or ≥3 days with ≥10h wear time^21^ were suggested. More recently, Buekers et al. recommended >12 h/day across ≥3 days during waking hours (7:00-22:00) for studies assessing walking activity and gait parameters^69^. However, as this recommendation was published only recently, it could not reasonably have guided the design of many studies included in this review. Therefore, the PROactive definition was used as the reference standard for the quality assessment; 13 studies met this requirement.

Although some studies predefined a set of criteria to decide which measurement periods can be included in the analysis^24,27,28,30–32,37,39,40,43,45,48,50,52,53^, quality-checks of the sensor outputs were not universally performed or reported (Table 2). Although established definitions for valid wear time exist, their application to the included studies was limited by insufficient reporting of wear time data, making it impossible to determine whether these criteria were met. Notwithstanding that inbuilt algorithms attempt to classify non-wear time reliably^71^, most devices can be taken off by the participants, making compliance still crucial in securing adequate measurement periods. Hence, the reported PA values may thus underestimate PA, as activity profiles might have been captured incompletely. Non-wear-related gaps reduce the validity of daily activity estimates and may also affect the reliability of measurements across days. Missing segments of activity data may therefore obscure relevant changes in DLPA. This limitation is particularly relevant in PAH populations, where daily fluctuations in symptoms and exertional capacity are described well. This has also been confirmed by a recently published analysis of accelerometry data from the PHANTOM trial (a randomized clinical trial with anastrozole in PAH)^72^.

Limitations of our review include the heterogeneity of the cohort resulting from the inclusion of PH subgroups, as PAH-SSc or CTEPH. Furthermore, potential DLPA-affecting comorbidities were not considered. Due to the heterogeneity of DLPA outcomes, the calculation of meta-estimates was only possible for a small number of outcomes or their correlation with traditional outcome measures in PH. Despite not using a conventional risk-of-bias assessment, restricting the review to peer-reviewed original research provided a minimum quality threshold; we appraised the conduct and reporting quality of wearable-based measurements using predefined criteria. This allowed concentrating the evaluation on the implementation and validity of DLPA parameters rather than on general study quality.

To the best of our knowledge, this is the first comprehensive systematic review and meta-analysis of digital assessment of DLPA in PH. We conclude that DLPA outcomes quantify patients’ actual behavior in real-world settings and seem to reflect a construct that is related to, yet distinct from, exercise capacity (6MWD), clinician-assessed disease severity (WHO-FC), and patient-reported activity experience. However, robust validation of these outcomes against established clinical is lacking in the current literature. The need for more rigorous and standardized assessments of DLPA (e.g., reporting of wear and non-wear), pre-defined follow-up examinations and pre-defined sets of parameters, is evident. The field now requires prospective studies and longitudinal validation of the responsiveness and prognostic value of DLPA outcomes.

## Data Availability

All data produced in the present work are available upon reasonable request to the authors

## Acknowledgement

SB: First reviewer, draft of manuscript, design of figures, quality assessment

LT: Statistical analysis (meta-analysis), data analysis and interpretation, quality assessment, draft of manuscript

YM: Second reviewer

CLR, AG, MB and CPJ: involved in the drafting of the manuscript and critically revising it for content

SUJ: Study design, third reviewer, data analysis and interpretation, writing of manuscript, takes responsibility for the integrity of the data and the accuracy of the data analysis

All authors have read and approved the final manuscript.

## Abbreviation List

6MWD: 6-minute walk distance
6MWT: 6-minute walk test
CAMPHOR: Cambridge pulmonary hypertension outcome review
CPET: Cardiopulmonary exercise testing
CTEPH: Chronic thromboembolic pulmonary hypertension
DLPA: Daily life physical activity
MET: Metabolic equivalent of task
NT-proBNP: N-terminal pro-B-type natriuretic peptide
NYHA FC: New York Heart Association functional class
PAH: Pulmonary arterial hypertension
PH: Pulmonary hypertension
WHO-FC: WHO functional class

