## Supplement for "Digital Assessment of Real-World Physical Activity in Pulmonary Hypertension: A Systematic Review and Meta-Analysis"

**Supplementary file**

**Detailed search strategy**

**PubMed**

| | | Search terms | |
|---|---|---|---|
| | **Pulmonary hypertension** | "hypertension, pulmonary"[MeSH Terms]<br>"pulmonary arterial hypertension"[All Fields]<br>"chronic thromboembolic pulmonary hypertension"[All Fields]<br>"pulmonary hypertension"[All Fields]) | OR |
| **AND** | **Physical activity** | "Exercise"[MeSH Terms]<br>"physical activity"[All Fields]<br>"mobil*"[All Fields]<br>"daytime activity"[All Fields]<br>"step*"[All Fields]<br>"walk*"[All Fields]<br>"exercise capacity"[All Fields] | OR |
| **AND** | **Wearable device** | "Wearable Electronic Devices"[MeSH Terms]<br>"Accelerometry"[MeSH Terms]<br>"Actigraphy"[MeSH Terms]<br>"actigraph*"[All Fields]<br>"activity tracker"[All Fields]<br>"activity monitor"[All Fields]<br>"wearable device"[All Fields]<br>"mobile health"[All Fields]<br>"smartwatch"[All Fields]<br>"Fitbit"[All Fields]<br>"track*"[All Fields]<br>"acceleromet*"[All Fields]<br>"pedometer"[All Fields]<br>"digital measurement"[All Fields] | OR |

*Table 3. Search strategy for Pubmed*

**Embase**

| | | Search terms | |
|---|---|---|---|
| | **Pulmonary hypertension** | 'pulmonary hypertension'<br>'pulmonary arterial hypertension'<br>'pulmonary hypertension'/exp | OR |
| **AND** | **Physical activity** | Exercise<br>'physical activity'<br>mobil*<br>'daytime activity'<br>step*<br>walk*<br>'exercise capacity' | OR |
| **AND** | **Wearable device** | 'wearable electronic devices'<br>Accelerometry<br>actigraph*<br>'activity tracker'<br>'activity monitor'<br>wearable device'<br>'mobile health'<br>smartwatch<br>'fitbit'<br>track*<br>'acceleromet*'<br>pedometer<br>'actigraph*'<br>'digital mesurement' | OR |
| | | 'Article'/it<br>'Article in Press'/it<br>'Preprint'/it | OR |

*Table 4. Search strategy for Embase*

### Devices and wear locations

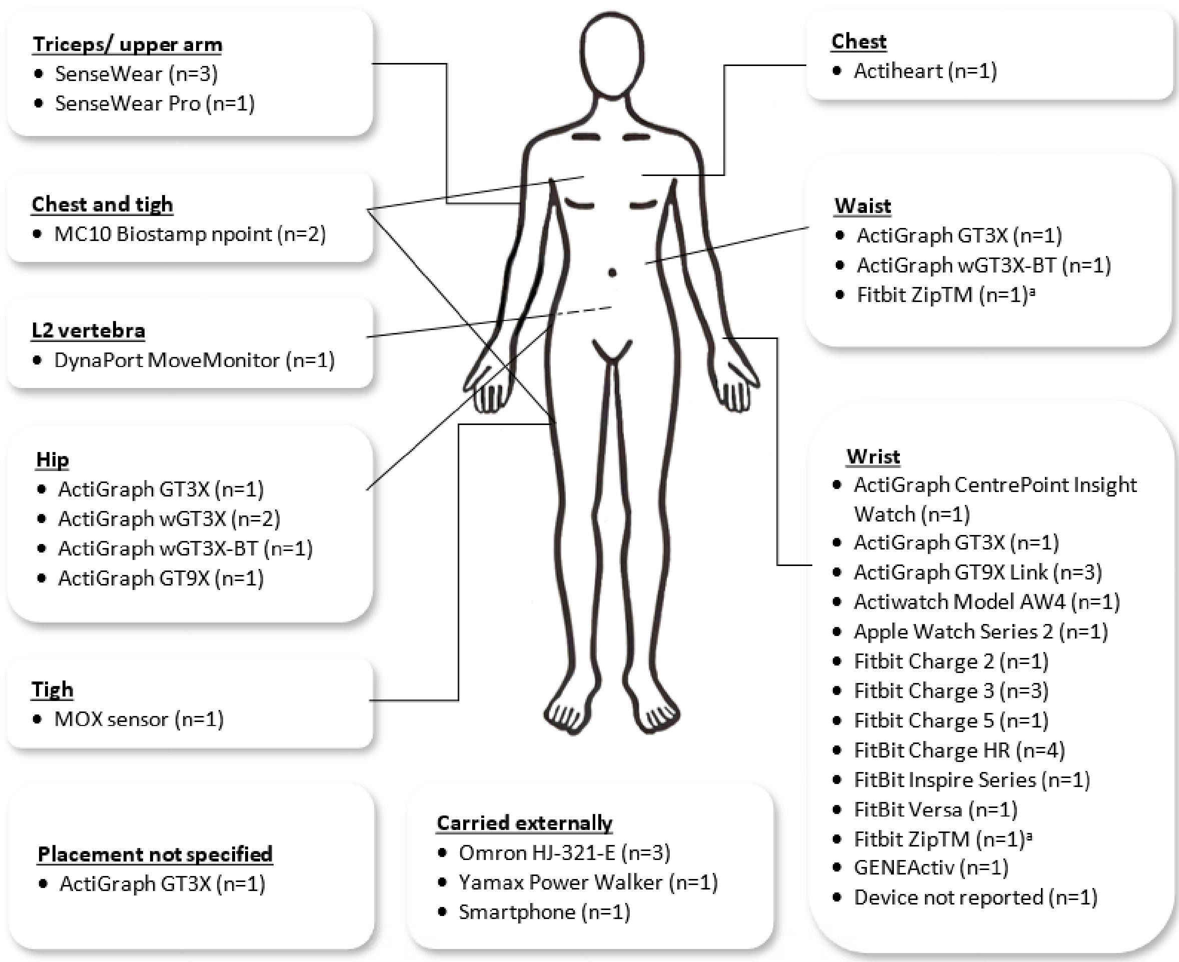


*Figure 4. Devices and their wear location. Illustration generated with DALL-E AI and modified manually; a = Device used in the same study but with different wear locations; n > number of studies, as some studies reported multiple device models.*

| First Author (year) | Group | DMOs and Six-minute-walk distance (6MWD) | DMOs and clinical functioning | DMOs and Patient Reported Outcomes (PROs) |
|---|---|---|---|---|
| Cascino (2019) | All participants | | | **↓ Daily steps ↑ Lack of interest in exercise***<br>**↓ Daily steps ↑ Lack of enjoyment from exercise***<br>**↓ Daily steps ↑ Lack of skills for exercise*** |
| González-Saiz (2018) | PH | | **↑ Daily inactivity time ↓ BMI*** | |
| Howard (2023)/ Rehmann (2025) | All participants | Baseline/ Week 16/ Week 24<br>**Daily steps ↑ 6MWD***<br>**↑ Daily activity time (NSPA) ↑ 6MWD***<br>**↑ Daily activity time (MVPA) ↑ 6MWD***<br>**↑ Cadence ↑ 6MWD***<br>**↑ Bout distance ↑ 6MWD***<br>**↑ Bout duration ↑ 6MWD*** | | |
| Hughes (2025) | All participants | Baseline<br>**↑ Daily steps ↑ 6MWD***<br>**↑ Daily activity time (sedentary) ↑ 6MWD***<br>**↑ Daily activity time (light activity) ↑ 6MWD***<br>**↑ Daily activity time (MVPA)↑ 6MWD*** | | **↓ Daily steps ↑ MLHF total score***<br>**↓ Daily steps ↑ MLHF physical score***<br>**↓ Daily steps ↑ MLHF emotional score***<br>**↓ Daily activity time (MVPA) ↑ MLHF total score***<br>**↓ Daily activity time (MVPA) ↑ MLHF physical score*** |
| Lachant (2022) | All participants (Actigraph) | **↑ Daily activity time (light activity) ↑ 6MWD***<br>**↑ Daily activity time (moderate activity) ↑ 6MWD***<br>**↑ Daily activity time (total) ↑ 6MWD*** | **↑ Daily activity time (light activity) ↓ WHO-FC***<br>**↑ Daily activity time (moderate activity) ↓ WHO-FC***<br>**↑ Daily activity time (total) ↓ WHO-FC***<br>**↑ Daily activity time (light activity) ↓ REVEAL 2.0***<br>**↑ Daily activity time (moderate activity) ↓ REVEAL 2.0***<br>**↑ Daily activity time (total) ↓ REVEAL 2.0***<br>**↑ Daily activity time (light activity) ↓ Age*** | **↑ Daily activity time (light) ↑ Activity level (remote survey)***<br>**↑ Daily activity time (moderate) ↑ Activity level (remote survey)***<br>**↑ Daily activity time (total) ↑ Activity level (remote survey)***<br>**↑ Daily activity time (moderate) ↑ Estimated activity time (remote survey)***<br>**↑ Daily steps ↑ Estimated activity time (remote survey)***<br>**↑ Daily activity time (light) ↓ Estimated inactivity time (remote survey)***<br>**↑ Daily activity time (moderate) ↓ Estimated inactivity time (remote survey)***<br>**↑ Daily activity time (total) ↓ Estimated inactivity time (remote survey)*** |
| | All participants (MC 10) | **↑ Daily activity time (total) ↑ 6MWD*** | **↑ Daily activity time (total) ↓ REVEAL 2.0***<br>**↑ Daily activity time (total) ↓ Age*** | **↑ Daily activity time (total) ↑ Estimated activity time (remote survey)***<br>**↑ Daily steps↑ Estimated activity time (remote survey)***<br>**↑ Daily activity time (total) ↑ Estimated inactivity time (remote survey)*** |

*Table 5. Reported correlations of DMOs 6MWD, clinical functioning parameters and PROs. *statistically significant correlation p-value < 0,05, Δ change between baseline and follow-up; Abbreviations: 1-STST: One-minute sit-to-stand test, 6MWD: 6-minute-walk distance, CAMPHOR: Cambridge pulmonary hypertension outcome review, CPM: counts per minute, DMO: Digital mobility outcomes, EE: energy expenditure, EQ-5D: Health related quality of life, EQ-VAS: EuroQol visual analogue scale, HADS-D: Hospital anxiety and depression scale (depression), HRR: Heart rate reserve, HRV: Heart rate variation, MET: Metabolic equivalent of task, MLHF: Minnesota living with heart failure, MRADL: Manchester respiratory activities of daily living questionnaire, MVPA: moderate to vigorous physical activity, NSPA: Non-sedentary physical activity, NYHA-FC: New York Heart Association functional class, PH: Pulmonary hypertension, Q-StO2: Quadriceps tissue oxygen saturation, SF-36 GH: Short form-36 General health scale, SF-36 PF: Short form-36 Physical function scale, SF-36 PR: Short form-36 Physical role scale, SF-36 SF: Short form-36 Social function scale, SF-36 V: Short form-36 Vitaliy scale, STST = stand-to-sit transfer, VE/VO2: Ventilatory equivalent for oxygen, VMC: Vector magnitude count, WHO-FC: WHO-functional class.*

| First Author (year) | Group | DMOs and Six-minute-walk distance (6MWD) | DMOs and clinical functioning | DMOs and Patient Reported Outcomes (PROs) |
|---|---|---|---|---|
| Lachant (2023) | All participants (Actigraph) | **↑ Daily steps ↑ 6MWD***<br>**↑ Daily activity time (total) ↑ 6MWD***<br>**↑ Daily activity time (light activity) ↑ 6MWD***<br>**↑ Daily activity time (moderate activity) ↑ 6MWD***<br>**↑ Average peak 5min steps ↑ 6MWD*** | **↑ Daily steps ↓ Age***<br>**↑ Daily activity time (total) ↓ Age***<br>**↑ Daily activity time (light activity) ↓ Age***<br>**↑ Daily activity time (moderate activity) ↓ Age***<br>**↑ Average peak 5min steps ↓ Age***<br>**↑ Daily steps ↓ Reveal 2.0 Lite Score***<br>**↑ Daily activity time (total) ↓ Reveal 2.0 Lite Score***<br>**↑ Daily activity time (light activity) ↓ Reveal 2.0 Lite***<br>**↑ Daily activity time (moderate activity) ↓ Reveal 2.0 Lite***<br>**↑ Average peak 5min steps ↓ Reveal 2.0 Lite***<br>**↑ Daily steps ↓ WHO-FC***<br>**↑ Daily activity time (total) ↓ WHO-FC***<br>**↑ Daily activity time (light activity) ↓ WHO-FC***<br>**↑ Daily activity time (moderate activity) ↓ WHO-FC***<br>**↑ Average peak 5min steps ↓ WHO-FC*** | **↑ Daily steps ↓ Borg dyspnea score***<br>**↑ Daily steps ↓ Emphasis 10***<br>**↑ Daily activity time (total) ↓ Emphasis 10***<br>**↑ Daily activity time (moderate activity) ↓ Emphasis 10***<br>**↑ Average peak 5min steps ↓ Emphasis 10***<br>**↑ Changes in Average peak 5min steps ↓ Changes in Emphasis 10*** |
| | All participants (MC 10) | **↑ Daily steps ↑ 6MWD***<br>**↑ Daily activity time (total) ↑ 6MWD*** | **↑ Daily steps ↓ Age***<br>**↑ Daily activity time (total) ↓ Age***<br>**↑ Daily steps ↓ Reveal 2.0 Lite Score***<br>**↑ Daily activity time (total) ↓ Reveal 2.0 Lite Score***<br>**↑ Daily steps ↓ WHO-FC***<br>**↑ Daily activity time (total) ↓ WHO-FC*** | **↑ Daily activity time (total) ↓ Borg dyspnea score*** |
| Lachant (2025) | | **↑ Daily steps ↑ 6MWD***<br>**↑ Peak steps ↑ 6MWD*** | | **↑ Daily steps ↓ Emphasis 10***<br>**↑ Peak steps ↓ Emphasis 10*** |
| Mainguy (2011) | PH Group | **↑ Daily steps ↑ 6MWD***<br>**↑ Daily EE > 3 METS (=MVPA) ↑ 6MWD***<br>**↑ Daily duration > 3 METs (=MVPA) ↑ 6MWD*** | **↑ Daily steps ↓ NYHA-FC***<br>**↑ Daily EE > 3 METS ↓ NYHA-FC***<br>**↑ Daily duration > 3 METs ↓ NYHA-FC*** | |
| Marvin-Peek (2021) | All participants | | **↑ Daily steps ↓ WHO-FC worsening at FU***<br>**↑ Daily steps ↓ Hospitalization*** | |
| Matura (2016) | All participants | **↑ Activity bouts ↑ 6MWD*** | **↓ Day-to-day physical activity variability ↑ Age*** | **↓ Daily CPM ↓ Energy subscale (US CAMPHOR)***<br>**↓ Day-to-Day Physical Activity Variability ↑ Mental Fatigue (MFI)***<br>**↓ % of activity bouts ↑ Total symptom subscale (US CAMPHOR)***<br>**↓ % of activity bouts ↓ Energy subscale (US CAMPHOR)*** |
| Minhas (2022) | Phantom | **↑ Daily steps ↑ 6MWD***<br>**↑ VMC ↑ 6MWD*** | | **↑ Daily steps ↓ emPHasis-10***<br>**↑ VMC ↓ emPHasis-10*** |
| | Penn-Cohort | **↑ Activity level cluster ↑ 6MWD*** | | |

*Table 5 – continued.*

| First Author (year) | Group | DMOs and Six-minute-walk distance (6MWD) | DMOs and clinical functioning | DMOs and Patient Reported Outcomes (PROs) |
|---|---|---|---|---|
| Nakazato (2021) | All participants | **↑ Daily steps ↑ 6MWD***<br>**↑ Activity time ↑ 6MWD*** | **↑ Activity time ↑ 1-STST***<br>**↑ Daily steps ↑ 1-STST*** | **↑ Daily steps ↑ MRADL***<br>**↑ Daily steps ↓ HADS-D***<br>**↑ Activity time ↑ MRADL*** |
| Okumus (2016) | All participants | **↑ Daily steps ↑ 6MWD***<br>**↑ Total EE ↑ 6MWD***<br>**↑ Active EE ↑ 6MWD*** | | **↑ Daily steps ↑ SF36 PF, PR, GH, V, MH, SF***<br>**↑ Daily steps ↑ MLHFQ***<br>**↑ EE ↑ SF36 PR, V, MH***<br>**↑ EE ↑ MLHF***<br>**↑ Active EE ↑ SF36 PF, GH, V, MH, SF*** |
| Panagiotou (2017) | All participants | **↑ Daily walking intensity (m/s) ↑ 6MWD*** | **↓ Daily walking intensity ↑ Age***<br>**↑ Daily walking intensity ↓ VE/VO$_2$***<br>**↑ Daily walking intensity ↑ TLCO %predicted***<br>**↑ Daily walking intensity ↑ HRR***<br>**↑ Daily walking intensity ↑ Q-StO$_2$ activity (%)***<br>**↑ Daily walking intensity ↑ Q-ΔStO$_2$ (%)*** | |
| Ploegstra (2025) | All participants | **↑ CPM ↑ 6MWD***<br>**↑ %MVPA ↑ 6MWD*** | **↑ CPM ↓WHO-FC***<br>**%MVPA ↓ WHO-FC*** | |
| Pugh (2012) | PH | **↑ Total activity count ↑ 6MWD*** | **↓ Total activity count ↑ WHO-FC***<br>**↑ Sedentary time ↑ WHO-FC*** | |
| Rosenzweig (2023) | All participants | <u>Follow-up</u><br>**↑ Daily steps ↑ 6MWD*** | | <u>Follow-up</u><br>**Δ ↑ Daily steps ↑ SF-36 PR*** |
| Saxer (2019) | All participants | <u>At rest</u><br>**↑ Daily steps ↑ 6MWD***<br>**↑ EE ↑ 6MWD***<br>**↑ Active EE ↑ 6MWD***<br>**↑ Daily activity 3 to 6 METs (=MVPA) ↑ 6MWD*** | | |
| Schmidt (2022) | All participants | **↑ MVPA (min/week) ↑ 6MWD*** | | **↑ MVPA (min/week) ↓ Symptoms dimension (CAMPHOR)*** |
| Sehgal (2019) | All participants | <u>Baseline</u><br>**↑ Daily steps ↑ 6MWD***<br><u>Follow-up change</u><br>**↑ Δ Daily steps ↑ 6MWD*** | | <u>Follow-up change</u><br>**↑ Moderately active minutes ↑ Δ EQ-VAS score***<br>**↑ Lightly active minutes ↑ Δ EQ-VAS score***<br>**↓ Sedentary minutes ↑ Δ EQ-VAS score***<br>**↑ Moderately active minutes ↑ Δ EQ-5D index***<br>**↑ Lightly active minutes ↑ Δ EQ-5D index***<br>**↓ Sedentary minutes ↑ Δ EQ-5D index*** |
| Sun (2025) | PH | **↑ HRV (Actiheart) ↑ 6MWD*** | | |
| Ulrich (2012) | All participants | | | **↑ Daytime activity counts/min ↓ SF-36 GH, V*** |
| Wieteska-Miłek (2022) | All participants | **↑ Daily steps ↑ 6MWD*** | | |
| Zijlstra (2017) | All participants | **↑ Daily activity time (MVPA) ↑ 6MWD***<br>**↑ Daily activity time (moderate PA) ↑ 6MWD*** | **↑ Daily activity time (MVPA) ↓ WHO-FC***<br>**↑ Daily activity time (moderate PA) ↓ WHO-FC***<br>**↑ Daily activity time (vigorous PA) ↓ WHO-FC*** | |

*Table 5 – continued.*

| First Author (year) | Group | DMOs and hemodynamic/imaging parameters | DMOs and laboratory parameters |
|---|---|---|---|
| González-Saiz (2018) | PH | **↑ Daily activity time (moderate activity) ↓ Mean arterial pulmonary pressure** | |
| Lachant (2022) | All participants (Actigraph) | | **↑ Daily activity time (light activity) ↓ NT-pro-BNP***<br>**↑ Daily activity time (total) ↓ NT-pro-BNP*** |
| | All participants (MC 10) | | **↑ Daily activity time (total) ↓ NT-pro-BNP*** |
| Lachant (2023) | All participants (Actigraph) | **↑ Daily steps ↓ Cardiac effort***<br>**↑ Daily activity time (total) ↓ Cardiac effort***<br>**↑ Daily activity time (light activity) ↓ Cardiac effort***<br>**↑ Daily activity time (moderate activity) ↓ Cardiac effort***<br>**↑ Average peak 5min steps ↓ Cardiac effort***<br>**↑ Daily steps ↑ Cardiac index***<br>**↑ Daily activity time (total) ↑ Cardiac index***<br>**↑ Daily activity time (light activity) ↑ Cardiac index***<br>**↑ Daily activity time (moderate activity) ↑ Cardiac index*** | **↑ Daily steps ↓ NT-pro-BNP***<br>**↑ Daily activity time (light activity) ↓ NT-pro-BNP***<br>**↑ Daily activity time (moderate activity) ↓ NT-pro-BNP***<br>**↑ Daily activity time (total) ↓ NT-pro-BNP***<br>**↑ Average peak 5min steps ↓ NT-pro-BNP***<br>**↑ Daily steps ↑ Hemoglobin*** |
| | All participants (MC 10) | **↑ Daily activity time (total) ↓ Cardiac effort***<br>**↑ Daily steps ↑ Cardiac index** | **↑ Daily steps ↓ NT-pro-BNP***<br>**↑ Daily activity time (total) ↓ NT-pro-BNP***<br>**↑ Daily steps ↑ Hemoglobin***<br>**↑ Daily activity time (total) ↑ Hemoglobin*** |
| Minhas (2022) | Phantom | **↑ VMC ↑ TAPSE***<br>**↑ VMC ↓ RV dysfunction*** | |
| | Penn-Cohort | | |
| Panagiotou (2017) | All participants | | **↑ Daily walking intensity ↓ log NT-proBNP*** |
| Ploegstra (2025) | All participants | | **↑ CPM ↓ NTproBNP*** |
| Saxer (2019) | All participants | <u>At rest</u><br>**↑ Daily steps ↑ SpO2***<br>**↑ EE ↑ SpO2***<br>**↑ EE ↓ Heart rate***<br>**↑ EE ↓ PAWP***<br>**↑ EE ↑ CO***<br>**↑ EE ↑ SV***<br>**↑ Active EE ↑ CO***<br>**↑ Active EE ↑ SV***<br>**↑ Daily activity 3 to 6 METs ↑ SpO2***<br>**↑ Daily activity 3 to 6 ↑ CO***<br>**↑ Daily activity 3 to 6 ↑ SV***<br><u>At maximal exercise</u><br>**↑ Daily steps ↑ Watts***<br>**↑ Daily steps ↑ Heart rate***<br>**↑ Daily steps ↑ CO***<br>**↑ Daily steps ↑ CO increase***<br>**↑ EE ↑ Watts***<br>**↑ EE ↓ PVR***<br>**↑ EE ↑ CO***<br>**↑ EE ↑ SV***<br>**↑ EE ↑ CO increase***<br>**↑ Active EE ↑ CO***<br>**↑ Daily activity 3 to 6 ↑ CO*** | |
| Ulrich (2012) | All participants | **↑ Time in bed ↓ PVR*** | |
| Woo (2022) | All participants | **↑ Daily steps ↓ TRV***<br>**↑ Sedentary time ↑ TRV*** | |

*Table 6 Reported correlations of DMOs with hemodynamic/ echocardiographic parameters and laboratory parameters. *statistically significant correlation p-value < 0,05. Abbreviations: CO: Cardiac output, CPM: counts per minute, DMO: Digital mobility outcomes, EE: energy expenditure, MET: Metabolic equivalent of task, NT-pro-BNP: N-terminal pro b-type natriuretic peptide, PH: Pulmonary hypertension, PVR: Pulmonary vascular resistance, RV: right ventricular, SpO2: Oxygen saturation, SV: Stroke volume, TAPSE: Tricuspid annular plane systolic excursion, TAPSV: Tricuspid annulus peak systolic velocity, TLCO: Transfer factor for carbon monoxide, TRV: Tricuspid regurgitation velocity, VMC: Vector magnitude count.*
